# Beyond staging: Potentially divergent trajectories of pain, quality of life, and fertility in endometriosis – prospective observational cohort study

**DOI:** 10.64898/2026.03.09.26347925

**Authors:** Michael Fanta, Zdeňka Lisá, Kristýna Hlinecká, Michal Mára, Radoslav Janoštiak

## Abstract

**Background:** Endometriosis is a heterogeneous disease in which anatomical lesion burden often shows poor correlation with pain severity and quality-of-life impairment. While classification systems such as the revised American Society for Reproductive Medicine (r-ASRM) and Enzian score accurately describe anatomical disease extent, their relationship to symptom burden and reproductive outcomes remains incompletely understood.

**Objective(s):** This study aimed to investigate the relationships between anatomical disease extent, pain severity, quality-of-life impairment, and fertility outcomes across ovarian, deep, and peritoneal endometriosis in a prospective cohort of women undergoing surgical treatment.

**Study Design:** This prospective observational cohort study included women aged 18–45 years undergoing laparoscopic surgery between 2023 and 2025 at a tertiary endometriosis center. Participants were categorized into ovarian (OE), deep (DE), or peritoneal (PE) endometriosis based on imaging and intraoperative findings. Pain severity was assessed using numeric rating scales across multiple domains, and quality of life was evaluated using the Endometriosis Health Profile (EHP-30+23). Anatomical disease burden was determined using r-ASRM and Enzian classifications. Patients were followed for 12 months after surgery to assess symptom trajectories, pregnancy outcomes, and surgical complications. A subset of lesion samples underwent RNA sequencing to explore molecular signatures associated with pain severity.

**Results:** A total of 145 women were included (OE n=33, DE n=55, PE n=25, controls n=32). Pain severity showed limited correlation with anatomical staging across subtypes. In contrast, infertility and the need for ureter surgery were strongly associated with higher Enzian scores and structural disease burden. Quality-of-life impairment closely paralleled pain intensity rather than anatomical stage. Transcriptomic analysis identified a molecular signature associated with high pain burden characterized by increased expression of inflammatory mediators *(IL6, CCL8, SPP1*), endocannabinoid system components (*PENK, CNR1)* and nociceptive transcription factors (*NR4A3, EGR3*). Longitudinal follow-up demonstrated substantial postoperative improvement in pain and quality of life independent of pregnancy outcomes.

**Conclusions:** Pain severity, quality-of-life impairment, and reproductive dysfunction in endometriosis represent partially independent dimensions of disease activity. While neuroinflammatory mechanisms appear to drive pain and quality-of-life impairment, fertility outcomes and organ-threatening complications are primarily determined by structural disease burden. Integrating anatomical staging with multidimensional symptom assessment and molecular profiling may enable more personalized management strategies for women with endometriosis.

## Introduction

Endometriosis is a chronic, inflammatory and estrogen-dependent disease affecting approximately 10–15% of women of reproductive age and up to 50% of women with infertility^1^. It is characterized by the presence of endometrium-like tissue outside the uterine cavity and presents with a highly heterogeneous clinical phenotype, ranging from asymptomatic disease to severe pelvic pain, organ dysfunction and impaired fertility^2^. The most frequently reported symptoms include dysmenorrhea, dyspareunia, chronic pelvic pain and bowel or urinary complaints, all of which may significantly reduce physical, sexual and professional activity and negatively impact quality of life^3^.

Despite decades of research, the relationship between anatomical disease extent and clinical symptom burden remains incompletely understood. The revised American Society for Reproductive Medicine (r-ASRM) classification has been widely used to stage endometriosis based on surgical findings^4^; however, multiple studies have demonstrated poor correlation between r-ASRM stage and pain severity^5,6^. Similarly, the Enzian and its updated version #Enzian were developed to provide a more detailed description of deep infiltrating endometriosis (DE) and compartment-specific involvement^7^. Although these systems improve and facilitate anatomical mapping and surgical planning, their correlation with patient-reported pain remains inconsistent^6,8^.

Recent prospective analyses have shown that the intensity of dysmenorrhea, dyspareunia and chronic pelvic pain does not systematically increase with more extensive compartment involvement in deep endometriosis^8,9^. Likewise, ultrasound-based Enzian (ENZIAN(u)) staging demonstrated that dysmenorrhea shows no clear compartment-specific pattern, while dyspareunia and bowel symptoms show only selective associations, primarily with retrocervical or uterosacral ligament involvement^8–10^. These findings support the concept that pain in endometriosis is multifactorial and not solely determined by lesion size or anatomical distribution.

In contrast, pathophysiology of fertility in endometriosis is very complex, and it appears more closely associated with anatomical disease burden. Warzecha et al. reported that infertility prevalence increases significantly with advanced r-ASRM stage, even though pain severity does not^11^. Severe disease was associated with prolonged time to conception and higher infertility rates, suggesting that reproductive prognosis depends more strongly on structural distortion, adhesions and deep compartment involvement than on symptom intensity^11^. This mismatch between anatomical extent and reproductive outcomes suggests that pain, quality-of-life impairment, and fertility may represent partially distinct (though interrelated) aspects of disease burden.

Furthermore, endometriosis is increasingly viewed as a multidimensional condition with systemic and psychosocial components. Chronic pelvic pain is associated with reduced quality of life and a higher prevalence of depressive symptoms, yet depressive symptom severity appears to correlate more strongly with pain burden than with disease stage^12–14^. These data highlight the limitation of purely lesion-based classifications and support the need for integrated clinical models that simultaneously consider anatomical, symptomatic and reproductive outcomes. Although prior studies have evaluated classification systems, symptom correlations, or quality-of-life impairment individually, few investigations have examined these domains concurrently across endometriosis subtypes or assessed their evolution longitudinally after laparoscopic surgical treatment. In particular, limited data exist comparing ovarian endometriosis (OE), deep endometriosis (DE), and peritoneal endometriosis (PE) with respect to pain trajectories, quality-of-life improvement, postoperative anatomical scores, and subsequent pregnancy outcomes within the same cohort.

Given the increasing emphasis on individualized management and fertility preservation strategies, a comprehensive analysis integrating anatomical staging, symptom burden, quality-of-life impairment and reproductive outcomes across endometriosis subtypes is warranted. Clarifying whether symptom improvement translates into improved fertility, and whether residual anatomical disease burden predicts reproductive success independent of pain, has direct implications for patient counseling and treatment planning.

The aim of the present study was therefore to investigate the relationships between (1) anatomical disease extent assessed by r-ASRM and Enzian classification, (2) pain severity and multidimensional quality-of-life impairment, and (3) postoperative pregnancy outcomes across ovarian, deep and peritoneal endometriosis. By integrating these domains in a longitudinal framework, we sought to clarify whether pain, quality of life and fertility represent convergent or independent axes of disease activity and to identify subtype-specific patterns that may inform personalized management strategies.

## Materials and Methods

### Study Design and Setting

This study was designed as a prospective observational cohort analysis with longitudinal follow-up. Women diagnosed with endometriosis were consecutively recruited at General University Hospital in Prague (2023-2025). The study included both cross-sectional baseline assessment and follow-up evaluation at 3 and 12 months after surgical intervention. The primary objective was to evaluate the relationship between anatomical disease burden, symptom severity, quality-of-life impairment, and reproductive outcomes across endometriosis subtypes. Secondary objectives included assessment of sterility status and predictors of ureter surgery.

The study was conducted in accordance with the Declaration of Helsinki and approved by the local Ethics committee of General University Hospital, Department of Gyneacology, Obstetrics and Neonatology (Ethical approval ID: Ref. No. 3/23 Grant GIP) on 16/02/2023. The recruitment of the patients for the study started on 01/04/2023. All participants provided written informed consent prior to inclusion.

### Participants

Inclusion criteria

- Women aged 18–45 years
- Availability of complete baseline pain and quality-of-life data
- Available r-ASRM and/or Enzian classification

Additional inclusion criteria – endometriosis group

- Laparoscopically and histologically confirmed diagnosis of endometriosis

Additional inclusion criteria – healthy controls

- No endometriosis detected intra-operatively or histologically
- Undergoing planned laparoscopic or hysteroscopic procedure for another benign indication

Exclusion criteria (both groups)

- Age <18 or >45 years
- Refusal to participate or withdrawal of consent
- Non-fulfilment of group-specific criteria at the time of surgery
- Active malignancy
- Severe systemic or psychiatric comorbidities that could independently affect quality of life
- Incomplete staging or missing follow-up data

Healthy controls were recruited among women undergoing gynecological laparoscopic surgery for other benign indications and had no clinical, imaging, or surgical evidence of endometriosis.

### Endometriosis Phenotyping

Participants were categorized into three subtypes based on dominant lesion type:

1. Ovarian endometriosis (OE)
2. Deep endometriosis (DE)
3. Peritoneal endometriosis (PE)

Subtype classification was determined preoperatively by expert sonographers using high-resolution transvaginal ultrasound according to the International Deep Endometriosis Analysis (IDEA) consensus criteria and intraoperatively by the operating surgeon.

### Anatomical Staging

Disease severity was assessed using:

- Revised American Society for Reproductive Medicine (r-ASRM) staging (I–IV)
- #Enzian classifciation - Enzian scores were recorded both preoperatively based on ultrasound (u) and intraoperatively based on surgical findings (s).)

Enzian compartments and severity grades were systematically documented as Enzian (u) by expert sonographers in the preoperative transvaginal ultrasound report and as surgical Enzian [Enzian (s)] by the operating surgeon in the operative report.

### Surgical Management

All surgeries were performed by experienced endometriosis surgeons within a multidisciplinary team. The surgical goal was complete excision of macroscopically visible lesions while preserving organ function and fertility when possible. Ureter surgery (e.g., ureterolysis or resection) was documented separately. Indications were based on intraoperative findings of ureteral involvement or obstruction.

### Pain Assessment

Pain was evaluated using an 11-point Numeric Rating Scale (NRS, 0–10) for the following domains:

- Chronic pelvic pain
- Dysmenorrhea (painful menstruation)
- Dyspareunia (painful sexual intercourse)
- Dysuria (painful urination)
- Dyschezia (painful defecation)

A composite pain score was calculated as the mean of individual pain domain scores. Pain assessment was performed:

- At baseline (preoperatively)
- At 3 months postoperatively
- At 12 months postoperatively

### Enzian score calculation

Enzian (u) and (s) scores were recorded by assigning compartment-specific severity grades (1–3) according to lesion size/extent for each affected compartment, including peritoneal (P), ovarian (O), tubal (T), and deep infiltrating compartments A, B, and C.

### Quality-of-Life Assessment

Quality of life was assessed using a structured questionnaire comprising four arbitrary domains:

- Physical and functional impairment
- Symptom burden and perceived control
- Emotional and psychological distress
- Social identity and stigma

The Endometriosis Health Profile-30+23 (EHP-30+23) was scored on a standardized scale; item scores were averaged to derive domain-specific and overall quality-of-life (QoL) impairment scores.

### Fertility and Reproductive Outcomes

Sterility status was defined as failure to conceive after ≥12 months of regular unprotected intercourse.

Pregnancy outcomes were assessed at 12-month follow-up. Both spontaneous conception and conception via assisted reproductive technologies (ART) were recorded.

Anti-Müllerian hormone (AMH) levels were documented where available, both preoperatively and at 3 months postoperatively.

### Longitudinal Follow-up

Patients were reassessed at:

- 3 months postoperatively
- 12 months postoperatively

The following were recorded at follow-up:

- Pain scores
- Quality-of-life (QoL) scores
- Clinical gynecological examination and Enzian (u) score
- Conception attempts and Pregnancy status

#### Pain-associated biomarker analysis

A subset of patient tissue samples included in this study was subjected to transcriptomic profiling to explore molecular signatures associated with pain severity and endometriosis subtype. RNA sequencing (RNA-seq) was performed as previously described^15^, with raw sequencing data deposited in the ArrayExpress database under accession number E-MTAB-15117. Sample selection was performed without clinical pre-stratification and was based solely on chronological procurement and fulfillment of predefined RNA quality criteria. The final cohort included samples from ovarian, deep, and peritoneal endometriosis lesions, as summarized in Supplementary Figure 1A. For exploratory analysis of pain-associated molecular signatures, patients were stratified into high-pain and low-pain groups based on composite pain scores derived from standardized numeric rating scale assessments at baseline. High-pain group corresponds to samples from women with composite pain score above median (>15.5), while low-pain group corresponds samples from women with composite pain score below median (<15.5). Pain stratification thresholds were predefined to ensure separation of clinically meaningful phenotypes while maintaining adequate statistical power. Gene expression was analyzed using standard bioinformatics pipeline in R (4.4.2) using Bioconductor package DEseq2^16^. Final p values were adjusted using the Benjamini and Hochberg method and genes with p-adjusted value <0.05 were considered significantly differentially expressed.

### Statistical Analysis

#### Between-group comparisons

Kruskal–Wallis tests were used for comparisons between groups followed by Dunn’s multiple comparisons test for comparison between individual groups. Fisher’s exact test was used for categorical variables.

#### Correlation analyses

Spearman correlation coefficients were calculated to assess relationships between:

- Pain scores and anatomical staging
- QoL scores and pain
- Fertility outcomes and anatomical parameters
- Longitudinal analysis

A p-value <0.05 was considered statistically significant. Statistical analyses were performed using Prism software (GraphPad).

## Results

### Patient characteristics

A total of 145 women were included: ovarian endometriosis (n = 33), deep endometriosis (n = 55), peritoneal endometriosis (n = 25), and healthy controls (n = 32). Age, BMI, age at menarche, and cycle length were comparable across groups. Pain profiles differed by phenotype: dysmenorrhea was most pronounced in deep endometriosis, while non-cyclic pelvic pain and dyspareunia were highest in peritoneal disease. Dyschezia was markedly elevated in deep and peritoneal endometriosis compared to ovarian disease and controls. Infertility was substantially more frequent in deep endometriosis (62%) than in ovarian (27%) or peritoneal endometriosis (32%), and lowest among controls (22%). Deep endometriosis was predominantly associated with advanced r-ASRM stage IV disease and extensive multi-compartment involvement on Enzian classification, whereas ovarian and peritoneal phenotypes were characterized by more localized disease patterns.

### Pain burden differs by subtype and demonstrates distinct internal structure

Pain scores for pelvic pain, dysmenorrhea, dyspareunia, dysuria, and dyschezia are shown in Figure 1A. Pain scores for pelvic pain, dysmenorrhea, dyspareunia and dyschezia were significantly different among the groups, while dysuria pain scores were not (**Fig. 1A**). The most prominent difference was in pain score for dysmenorrhea where patients with all endometriosis subtypes reported significantly higher pain scores compared to healthy control. In other individual pain scores, there were apparent differences between subtypes. Women with peritoneal endometriosis reported significantly higher pain scores across all pain domains (except dysuria), while women with deep endometriosis reported higher pain scores for pelvic pain and for dyschezia with even higher magnitude than women with peritoneal endometriosis. Correlation heatmaps (**Fig. 1B**) revealed subtype-specific pain architecture. In OE, correlations between domains were generally weak to moderate, with the strongest association observed between pelvic pain and dyspareunia (**Fig. 1B, top left**). Similarly in DE, correlations remained modest with strongest correlation between dyspareunia, dysuria and dyschezia (**Fig. 1B, top right**). In contrast, PE exhibited strong positive correlations across most pain domains, indicating a more globally coherent pain phenotype (**Fig. 1B, bottom left**). Healthy controls demonstrated moderate inter-domain correlations at lower absolute pain levels (**Fig. 1B, bottom right**).

**Figure 1.**
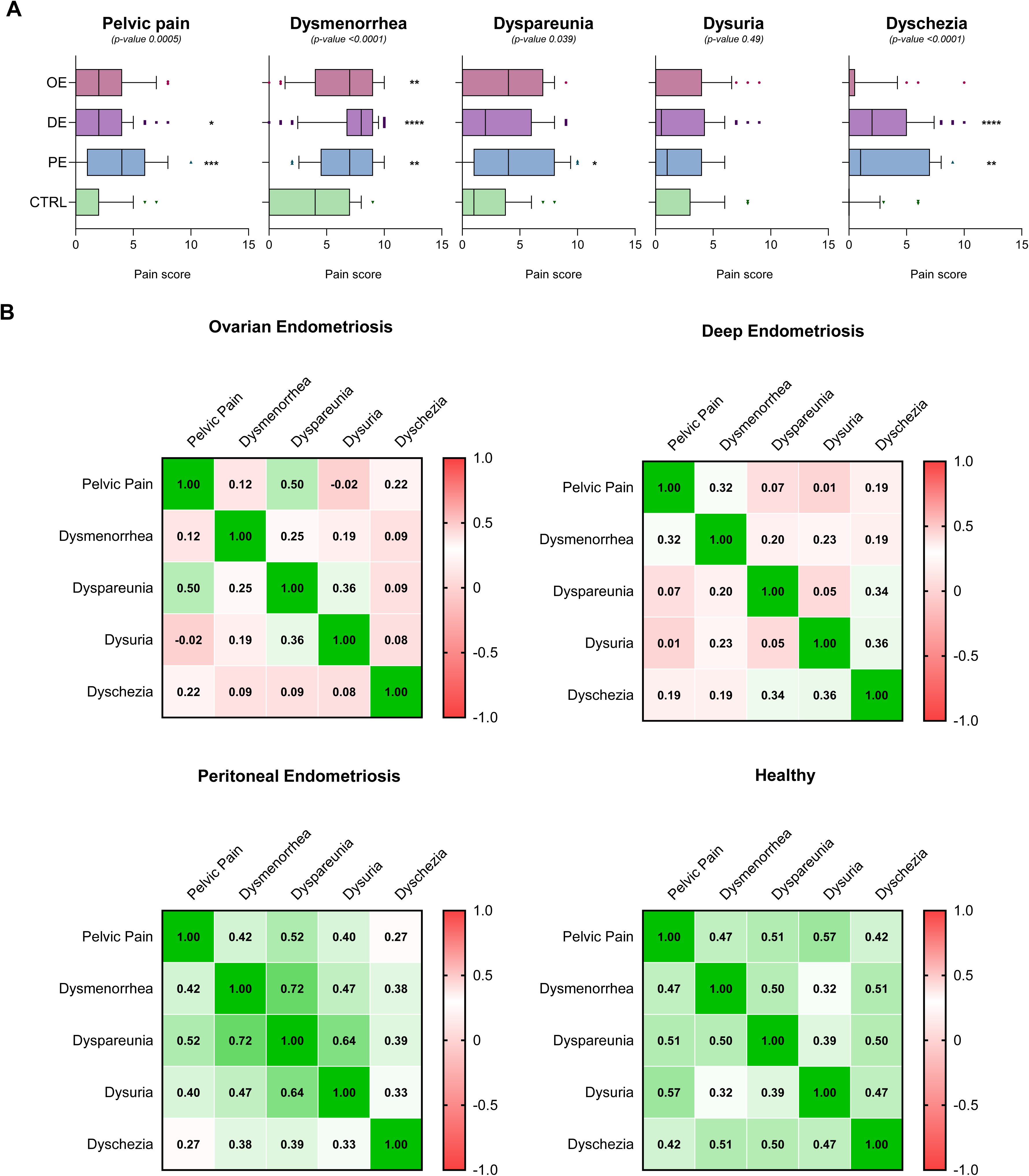
Pain score distribution and inter-domain correlations. (A) Boxplots showing pain scores for pelvic pain, dysmenorrhea, dyspareunia, dysuria and dyschezia in ovarian endometriosis (OE), deep endometriosis (DE), peritoneal endometriosis (PE), and healthy controls (CTRL). P-value represents Kruskal-Wallis test results, and asterisks represent Dunn’s test scores between control group and individual endometriosis subtypes as follows: * <0.05, ** <0.01, *** <0.001, **** <0.0001. (B) Spearman correlation heatmaps illustrating relationships between pain domains within each group.

### Pain shows limited correlation with anatomical staging and imaging findings

Figure 2 illustrates correlation matrices between pain scores and ultrasound and laparoscopic staging parameters across OE, DE, and PE. The correlation heatmap allows to draw several conclusions. In OE and DE, the pain scores do not correlated with Enzian scores (ultrasound or laparoscopic) or with r-ASRM stage, while in PE, we can see a correlation between individual pain scores and Enzian compartment A and B involvement assessed using ultrasound. The dissociation was most evident in OE and DE, where high anatomical burden did not correspond to proportional increases in symptom severity. Interestingly, there are differences in scoring correlation between ultrasound and laparoscopic Enzian scores. In OE, there is significant concordance between ultrasound and laparoscopic Enzian scores for compartments A, B and C involvement as well as for ovarian involvement (**Fig 2A**). Similarly, in DE, strongest correlation is seen in ultrasound and laparoscopic Enzian score for ovarian and tube involvement, and weaker correlation for compartments A, B and C (**Fig 2B**). Finally, in PE, we see again the strongest correlation between ultrasound and laparoscopic Enzian scores for ovarian and tube involvement, and much weaker and inconsistent between scores for compartment A, B and C (Fig. 2C). When looking at the correlation between r-ASRM stage and other features, the results are fairly consistent between subtypes. There is a positive correlation between r-ASRM and laparoscopic Enzian scores for compartments A, B, C and tube involvement in all three subtypes, with strongest correlation in PE, followed by OE and DE (Fig. 2A**-C**). Similarly, there is positive correlation between between r-ASRM and laparoscopic Enzian scores for compartments A, B, C and tube involvement in PE and OE, while in DE the correlation is weak and not significant (Fig. 2A**-C**). Finally, we compared the symptom severity and Enzian score-informed disease extent with r-ASRM stages (Fig. 3). Ultrasound and laparoscopic Enzian scores increased progressively with advancing r-ASRM stage across OE and DE, confirming structural concordance between staging systems (Fig. 3A-B), while in PE, r-ASRM staging was correlated only with laparoscopy-informed Enzian score reflecting higher difficulty with ultrasound assessment of peritoneal endometriosis (Fig. 3C). In contrast, pain domains and composite pain score showed substantial overlap across stages without consistent escalation.

**Figure 2.**
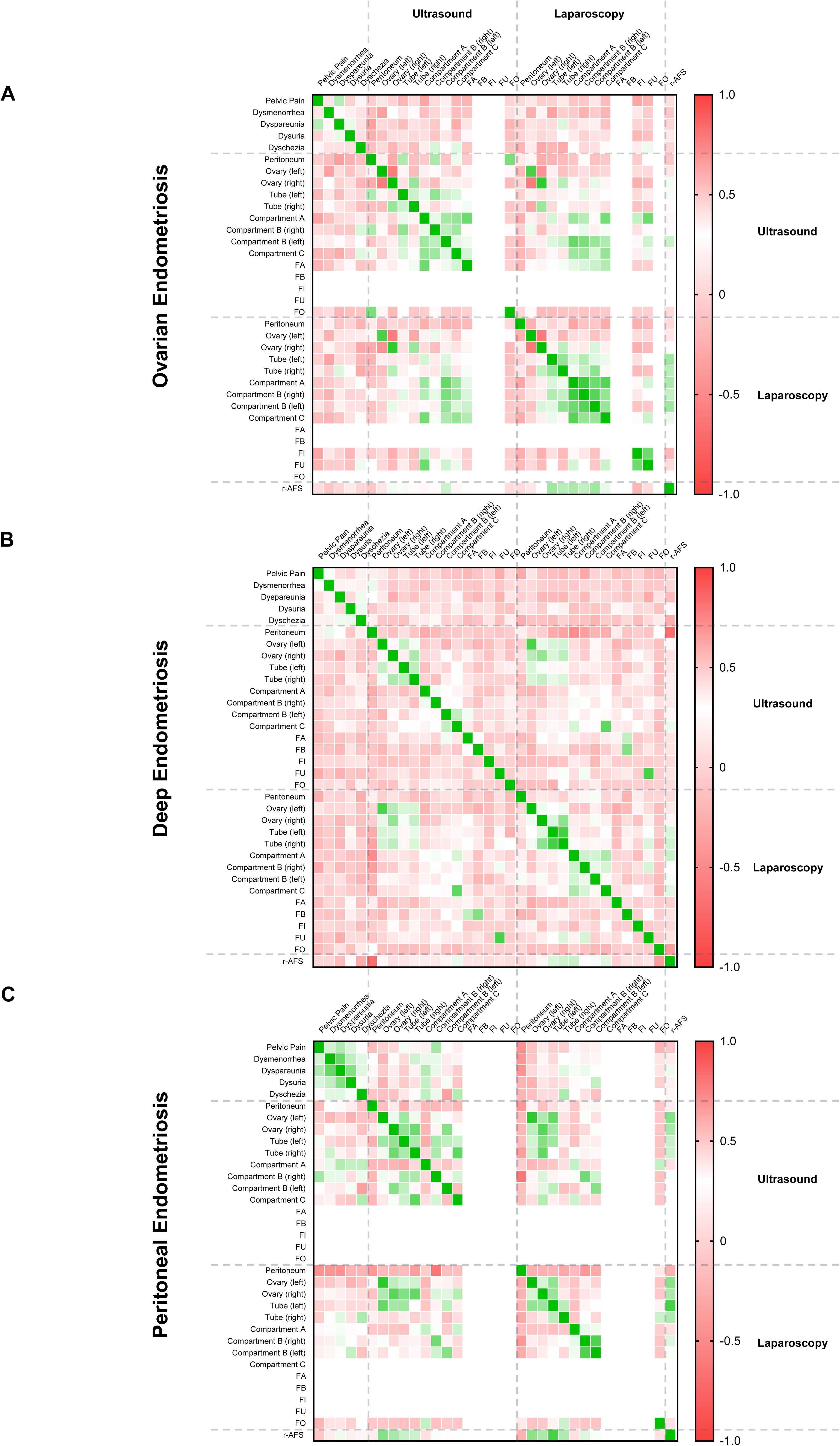
Correlation between pain, ultrasound, and laparoscopic findings. (A-C) Spearman correlation matrices for OE, DE, and PE demonstrating relationships between pain scores and ultrasound and laparoscopic Enzian parameters, including compartmental involvement and r-ASRM stage.

**Figure 3.**
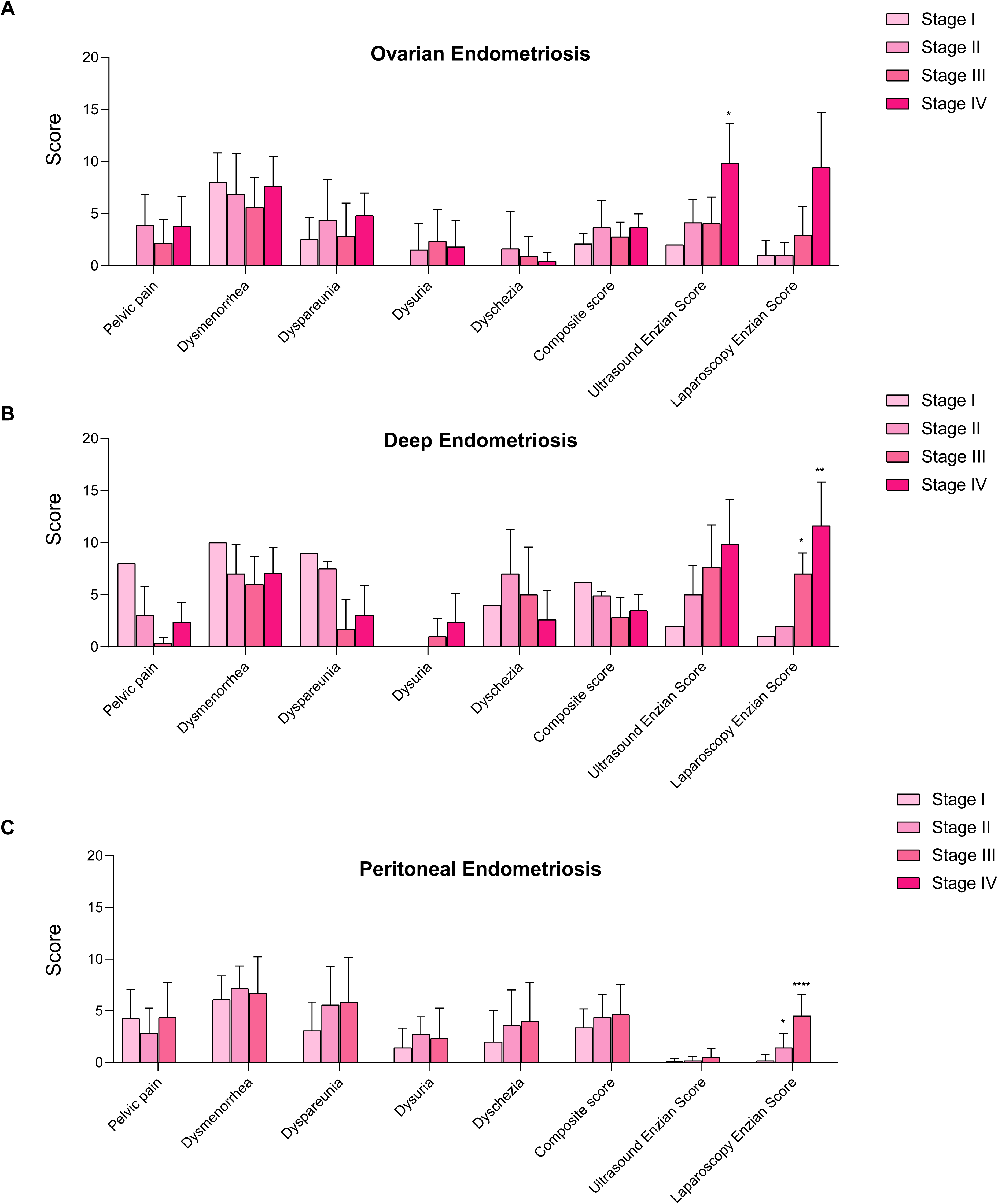
Pain and structural scores stratified by r-ASRM stage. (A-C) Bar graphs showing pain domain scores, composite pain score, ultrasound Enzian score, and laparoscopic Enzian score across r-ASRM stages I–IV within OE, DE, and PE. Asterisks represent Student’s t-test scores between control group and individual endometriosis subtypes as follows: * <0.05, ** <0.01, *** <0.001, **** <0.0001.

### Quality of life impairment is primarily pain-driven

Quality-of-life domain scores are presented in Figure 4A. All endometriosis groups demonstrated significantly worse physical and functional impact, symptom burden, emotional distress, and social identity impairment compared with controls. Women with DE and PE reported significantly increased QoL impairment scores across all 4 domains, with symptom burden and perceived control impairment being the most significant (Fig. 4A). In contrast, women with OE reported modestly increased QoL burden across the domains compared to healthy individuals, with symptom burden and perceived control impairment being the only one significantly increased (Fig. 4A). Correlation heatmaps (Fig. 4B) showed notable variability between subtypes. The strongest correlation was observed between QoL impairment and dysmenorrhea in women with OE, while there was no correlation between QoL impairment and general pelvic pain or dyschezia, and only a weak and inconsistent correlation with dysuria (Fig. 4B). Similar to women with OE, women with DE reported significant correlation between QoL impairment and dysmenorrhea, but also high correlation with general pelvic pain and dyschezia to a certain degree (Fig. 4B). On the other hand, women with PE consistently reported strong correlation between QoL impairment across all domains and symptoms severity, with strongest correlation between dysmenorrhea and dyspareunia with physical impact and symptoms burden (Fig. 4B) To explore the association between pain severity and molecular alterations within endometriotic lesions, we reanalyzed RNA sequencing data as previously described^15^. Eligible samples were stratified into high- and low-pain groups based on composite pain scores, and differential gene expression analysis was performed. Despite the relatively small cohort size (high pain: n = 13; low pain: n = 13), we identified a distinct set of genes significantly overexpressed in lesions from women with high pain burden (**Supplementary Figure 1B; Fig. 4C**). Among the most strongly upregulated transcripts were several genes with established roles in inflammatory signaling and pain perception. Notably, *IL6, OSM, CCL8 and SPP1* were significantly increased, consistent with enhanced inflammatory cytokine signaling (**Fig. 4C, top row**). The immediate early transcription factors *NR4A3* and *EGR3* associated with nociception were also markedly elevated, suggesting increased transcriptional activation associated with nociceptive signaling (**Fig. 4C, bottom left**). Finally, upregulation of endocannabinoid system-associated genes *PENK* and *CNR1* further supports engagement of neurobiological pathways within high-pain lesions (**Fig 4C**).

**Figure 4.**
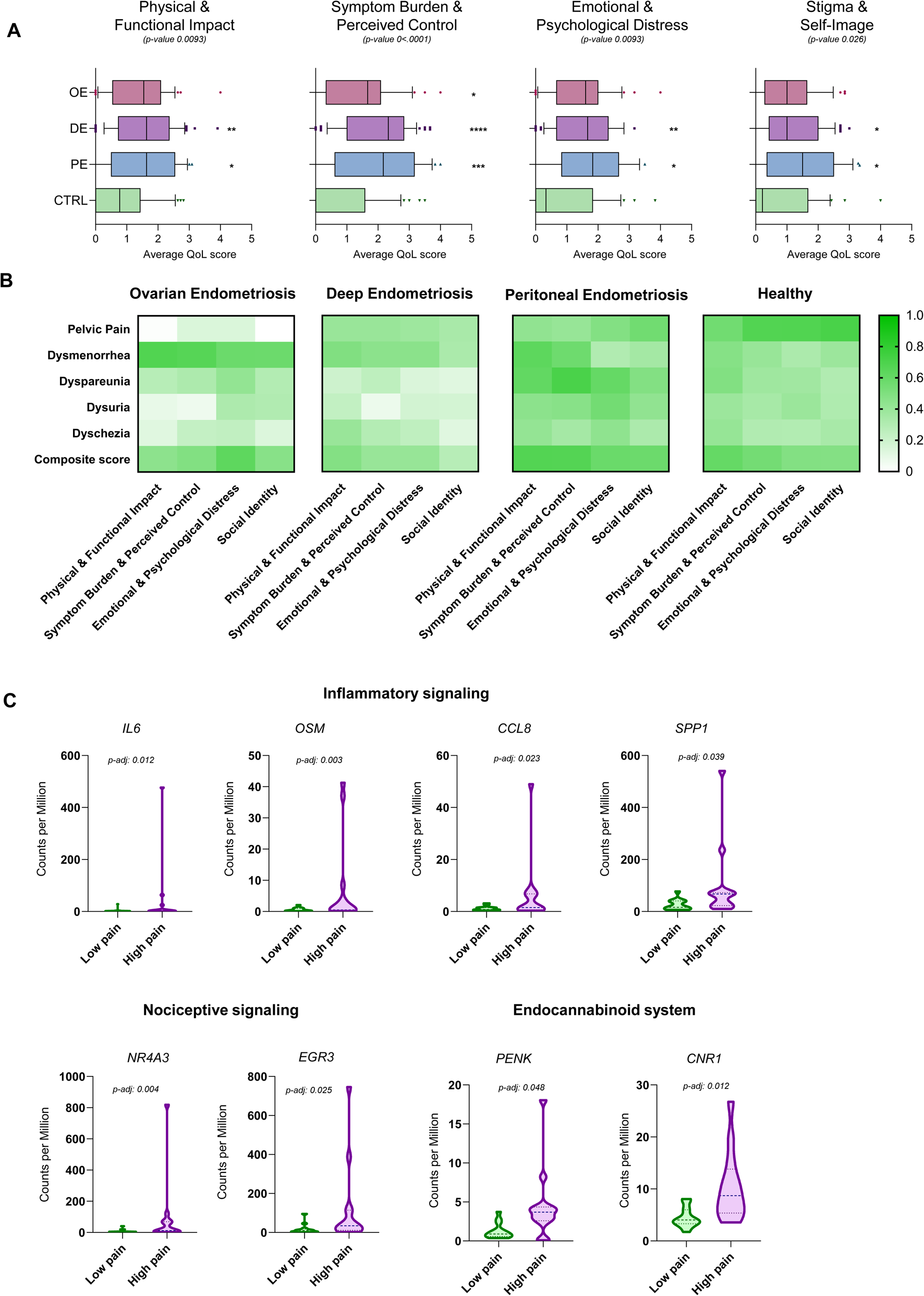
Quality of life and its association with pain. (A) Boxplots showing average Quality of life composite scores for 4 distinct domain, namely physical & functional impact, symptom burden & perceived control, emotional & psychological distress, stigma & self-image in ovarian endometriosis (OE), deep endometriosis (DE), peritoneal endometriosis (PE), and healthy controls (CTRL). P-value represents Kruskal-Wallis test results, and asterisks represent Dunn’s test scores between control group and individual endometriosis subtypes as follows: * <0.05, ** <0.01, *** <0.001, **** <0.0001. (B) Spearman correlation heatmaps illustrating relationships between pain domains QoL domains within each group. (C) Box plots showing counts per million (cpm) distribution for selected genes in high and low pain groups. P-value represents adjusted p-values calculated using Wald test followed by Benjamini and Hochberg adjustment.

### Infertility is associated with structural disease burden but not with pain

Infertility analyses are shown in Figure 5. Across OE, DE, and PE, women with confirmed infertility did not exhibit higher pain scores or composite pain burden compared with non-sterile women in the same subgroup (Fig. 5A**-C**). Similarly, AMH levels showed no consistent association with infertility status across the groups. In contrast, sterile women demonstrated consistently higher laparoscopic Enzian scores across subtypes, however reaching statistical significance only in women with DE (Fig. 5A**-C**). There was a trend showing higher ultrasound Enzian scores in women with confirmed infertility however not reaching statistical significance in any of the subtypes. On the other hand, r-ASRM stages were not correlated with infertility in women with OE and DE, while the women with PE with higher r-ASRM stages exhibited significantly higher proportion of infertility (Fig. 5C). These findings indicate that reproductive impairment is structurally mediated rather than symptom-driven.

**Figure 5.**
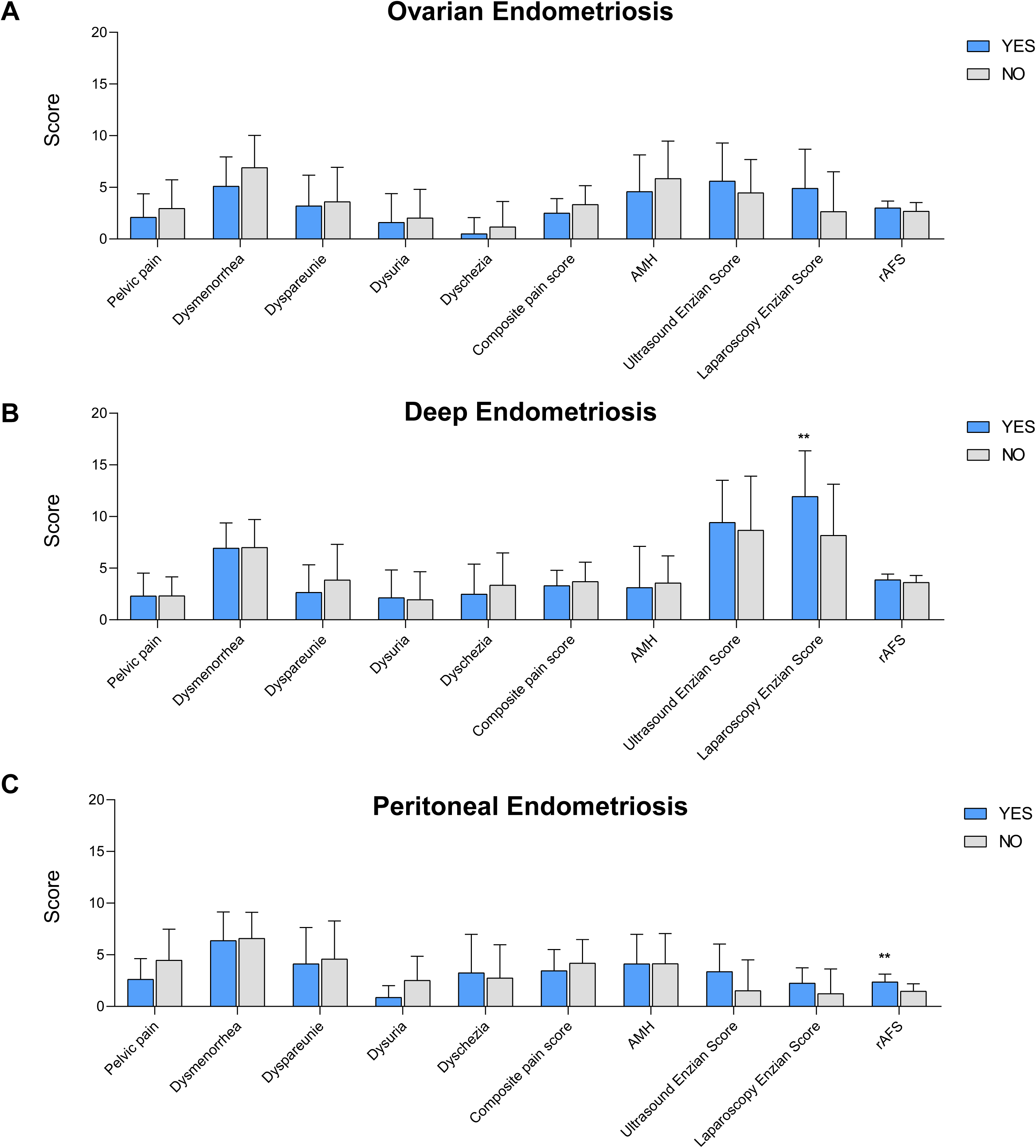
Association between sterility and clinical parameters. (A) Bar charts representing pain scores, AMH, ultrasound Enzian score, laparoscopic Enzian score, and r-ASRM stage in infertile (YES) versus non-infertile (NO) women across subtypes. Asterisks represent Student’s t-test scores between control group and individual endometriosis subtypes as follows: * <0.05, ** <0.01, *** <0.001, **** <0.0001.

### Longitudinal follow-up demonstrates symptom improvement independent of pregnancy

To investigate the functional, symptomatic and emotional recovery after laparoscopic removal of endometriotic lesions, we conducted a follow-up analysis at 3 and 12 months after surgery (Fig. 6A-C). Across all the subtypes, the trend was consistent with subtle differences. Pain perception across all the domains was most significantly decreased in women with higher r-ASRM (III/IV) staging at baseline with consistent decrease of follow up ultrasound Enzian scores (Fig. 6A). Interstingly, in women with lower r-ASRM stage (I/II) the pain perception decrease was rather modest or not evident, even though baseline pain scores were comparable high to women with higher stages and the follow up ultrasound Enzian score was lowered (Fig. 6A). On the other hand, quality-of-life scores improved consistently across subtypes and stages, further supporting a disconnection between symptoms, disease extent and quality of life scores (Fig. 6B). Additional analysis of reproductive health improvement showed that despite objective disease removal, pregnancy rates at 12 months did not significantly improved and pregnancy occurred in 6/11 women with OE, 9/30 with DE, and 4/10 with PE, without statistically significant differences across subtypes (Fig. 6C). Additional correlation analysis of pregnancy predictors showed overall poor correlation between recorded metrics (symptoms, stage, Enzian, QoL) at baseline or at 12 months follow up (**Supplementary Table 1**). Only statistical significant predictors were discovered in women with DE, where the pregnancy was associated with lower 12-month ultrasound Enzian score and significantly lower impairment in physical and functional impact, symptom burden, emotional distress, and social identity domains.

**Figure 6.**
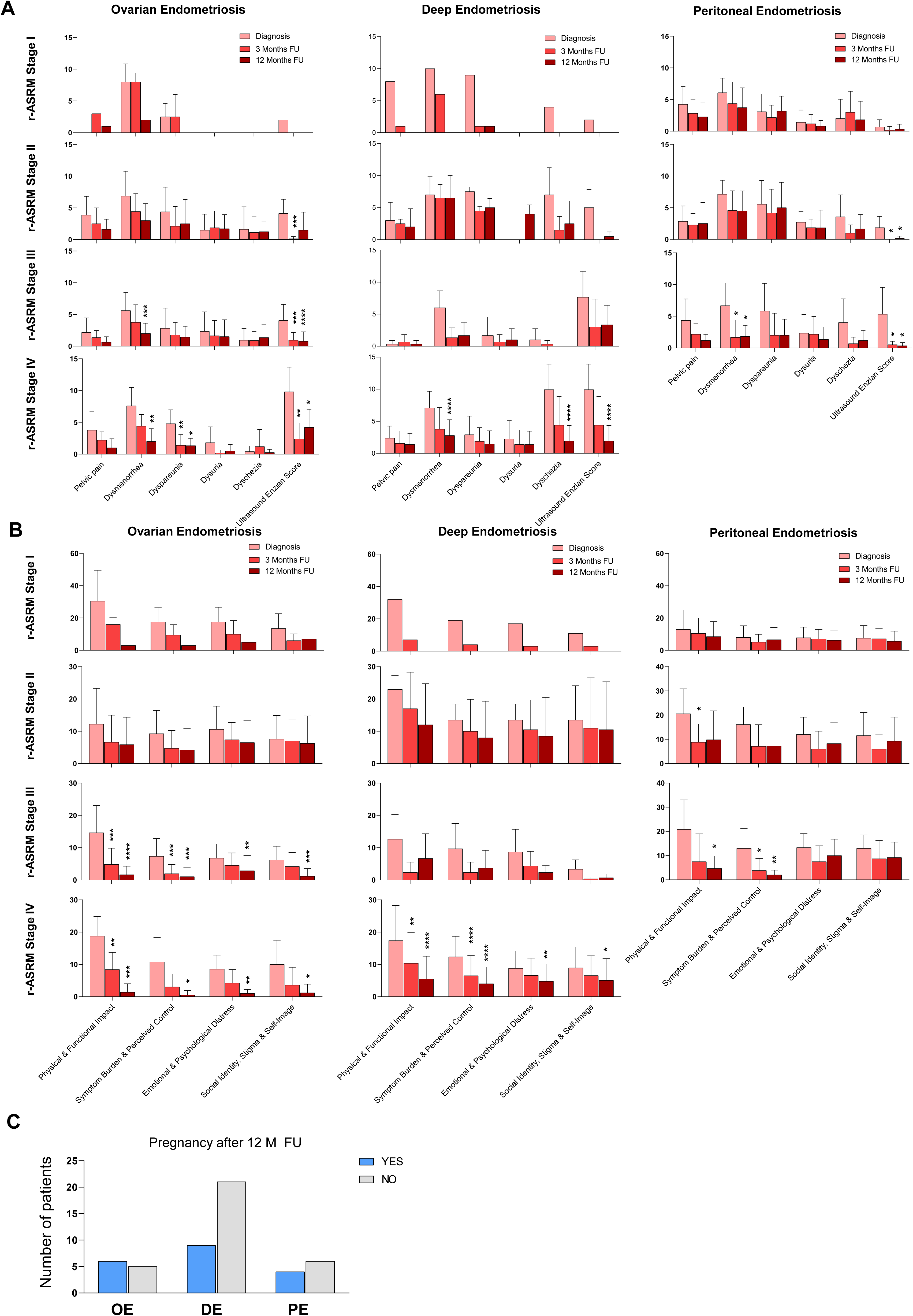
Follow-up trends and pregnancy outcomes. (A) Bar charts representing changes in pain scores and ultrasound Enzian score from diagnosis to 3 and 12 months follow up, grouped by rASRM stage and subtype. (B) Bar charts representing changes in QoL domain scores and ultrasound Enzian score from diagnosis to 3 and 12 months, follow up grouped by rASRM stage and subtype. (C) Bar charts representing number of pregnancies recorded at 12-month follow-up stratified by subtype. Asterisks represent Student’s t-test scores between control group and individual endometriosis subtypes as follows: * <0.05, ** <0.01, *** <0.001, **** <0.0001.

### Ureter surgery is driven by anatomical extent rather than symptoms

Since one of the major complication of deep endometriosis is possible renal failure, we investigated if there are any predictors of the necessity for ureter surgery. As shown in Figure 7, laparoscopic composite Enzian score as well as laparoscopic individual Enzian scores for tube involvement showed strong positive correlation for the need for ureter surgery, while right and left tubal involvement, compartment B lesions, ovarian involvement, showed significant yet much weaker association (Fig. 7A**-B**). Interestingly dyschezia and dyspareunia demonstrated medium negative associations but did not reach the statistical significance observed for structural parameters (Fig. 7A**-B**).

**Figure 7.**
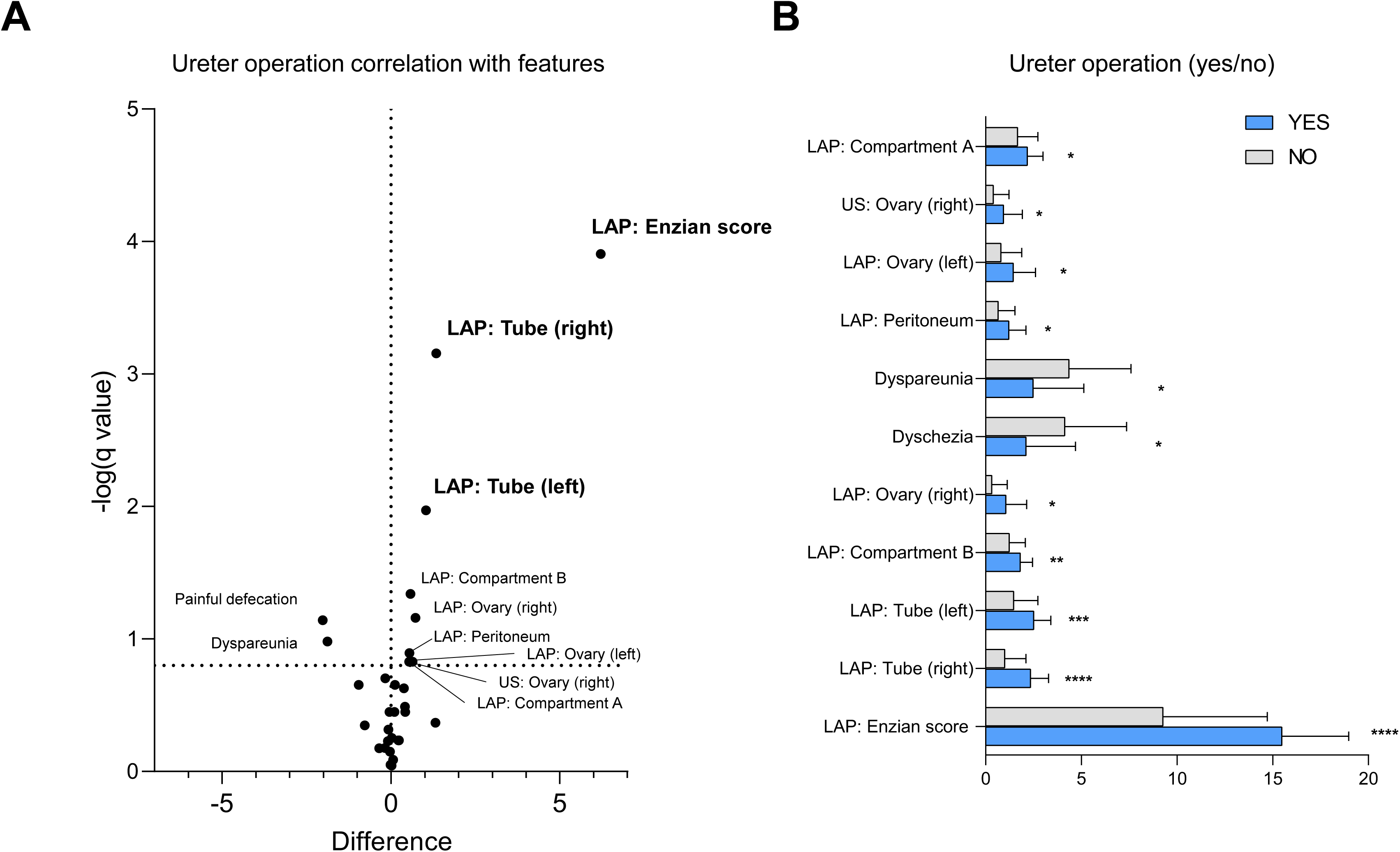
Ureter surgery outcomes. (A) Volcano plot showing characteristics associated with ureter surgery (difference vs –log(q value)). (B) Bar chart shows comparison of structural and symptom variables between patients with and without ureter surgery. Asterisks represent Student’s t-test scores between control group and individual endometriosis subtypes as follows: *<0.05, ** <0.01, *** <0.001, **** <0.0001. [LAP] – laparoscopy, [US] - ultrasound

## Comments

### Principal findings

In this study, we show that the clinical severity of endometriosis cannot be explained by a single anatomical continuum. Instead, our findings support the presence of two partially independent biological axes that shape disease manifestations: a neuroinflammatory axis governing pain severity and quality-of-life impairment, and a structurally driven fibrotic axis determining fertility outcomes, surgical complexity, and risk of organ compromise. Pain intensity and quality-of-life measures were largely independent of anatomical staging, whereas reproductive outcomes and surgical interventions were strongly associated with disease extent measured by r-ASRM and Enzian scores. Transcriptomic analysis further identified a molecular program associated with severe pain characterized by endocannabinoid system activation, inflammatory cytokine signaling, extracellular matrix remodeling, and activation of nociceptive transcriptional pathways. Together, these findings provide a biological framework that explains the longstanding discordance between lesion burden and symptom severity in endometriosis.

### Results

The weak association between anatomical staging and pain severity observed in our cohort is consistent with prior studies demonstrating poor correlation between r-ASRM stage and dysmenorrhea, chronic pelvic pain, or dyspareunia^8–10^. Pain in endometriosis arises from a complex interaction of inflammatory mediators, neuroangiogenesis, nerve fiber infiltration, and central sensitization. These mechanisms are not captured by lesion-based staging systems^17^. Fibrotic remodeling appears to be a central component of disease progression^18,19^. Deep infiltrating lesions are characterized by dense collagen deposition, myofibroblast activation, smooth muscle metaplasia, and extracellular matrix stiffening. These changes can mechanically stimulate nociceptors and alter local mechanotransduction^20,21^. Importantly, fibrotic remodeling does not necessarily correlate with total lesion burden. Small but highly fibrotic nodules may generate substantial mechanical tension and nerve entrapment, potentially explaining the heterogeneity of pain severity among patients with similar anatomical stages. Consistent with this model, transcriptomic analysis identified a distinct molecular signature associated with high pain burden across lesion subtypes. Lesions from patients with severe pain showed increased expression of inflammatory mediators including IL6, OSM, CCL8, and SPP1, suggesting a cytokine-driven microenvironment capable of directly sensitizing peripheral nociceptors through JAK/STAT and MAPK signaling pathways^22–24^. Elevated expression of the immediate early transcription factor NR4A3 and EGR3 further suggests activation of nociceptive transcriptional programs, as NR4A family members and EGR3 are rapidly induced in sensory neurons during inflammatory stimulation and contribute to persistent neuronal sensitization. Alongside this activation signal, we observed consistent upregulation of PENK, encoding the precursor of endogenous opioid peptides, and CNR1, encoding the cannabinoid receptor type 1. Both pathways are central components of intrinsic pain-modulatory systems and are commonly activated in response to chronic nociceptive stimulation^25,26^. Their upregulation therefore likely reflects a compensatory neuroregulatory response, in which endogenous opioid and endocannabinoid signaling attempts to counterbalance persistent inflammatory activation of sensory pathways. The simultaneous presence of inflammatory mediators and endogenous analgesic signaling suggests that painful lesions represent dynamic neuroimmune microenvironments characterized by both nociceptive activation and intrinsic attempts at pain modulation.

Quality-of-life impairment in our cohort closely paralleled pain severity and showed minimal association with anatomical staging. This observation is consistent with previous studies demonstrating that psychosocial distress and functional impairment in endometriosis are primarily mediated by chronic pain rather than lesion burden^6,11^. These results underscore the importance of incorporating validated patient-reported outcome measures into clinical evaluation. In contrast, fertility outcomes were strongly associated with anatomical disease burden. Higher Enzian scores and advanced r-ASRM stages were associated with sterility and reduced probability of pregnancy. This finding aligns with previous reports linking advanced endometriosis to infertility. Structural distortion caused by fibrosis and adhesions likely underlies this relationship. Progressive collagen deposition restricts fimbrial mobility, impairs tubal patency, and disrupts the spatial relationship between the ovary and fallopian tube necessary for efficient oocyte capture^27–29^.

Deep infiltrating endometriosis represents a clinical context in which structural and symptomatic axes may converge. In this subgroup, pregnancy within 12 months was associated with lower residual Enzian scores and improved quality-of-life measures. These findings suggest that complete surgical excision may alleviate both fibrotic mechanical distortion and sustained nociceptive signaling. However, the benefits of extensive surgical intervention must be balanced against potential risks, including repeated procedures and impacts on ovarian reserve. Ureteral involvement represents one of the most clinically significant manifestations of the structural axis. In our cohort, ureter surgery was strongly predicted by compartmental disease extent rather than symptom severity. Ureteral endometriosis often develops through extrinsic compression by parametrial or uterosacral fibrotic nodules (extrinsic) or by infiltration of ureteral wall (intrinsic form), leading to progressive luminal narrowing, impaired peristalsis, and hydronephrosis^30,31^. Because this process frequently progresses without prominent symptoms, reliance on pain severity alone may delay diagnosis. Chronic obstruction can result in hydronephrosis, renal cortical thinning, irreversible nephron loss, and ultimately renal failure. These findings highlight the importance of careful anatomical assessment and imaging surveillance in patients with deep compartment involvement.

### Clinical implications

These findings have several implications for clinical management. First, anatomical staging alone cannot reliably estimate symptom burden. Patients with limited visible disease may experience severe pain, whereas individuals with extensive lesions may remain relatively asymptomatic. Routine integration of validated patient-reported outcome measures and multidimensional pain assessment is therefore essential.

Second, the molecular characteristics associated with severe pain suggest that therapies targeting prostaglandin synthesis, such as nonsteroidal anti-inflammatory drugs, may be less effective in patients whose symptoms are driven predominantly by cytokine-mediated nociceptor sensitization and fibrotic tissue remodeling. Instead, the transcriptional profile observed in high-pain lesions—characterized by inflammatory mediators (IL6, OSM, CCL8, SPP1) and neuronal activation markers (NR4A3, EGR)—together with increased expression of endogenous pain-modulatory pathways including PENK and CNR1, suggests an active neuroimmune pain phenotype. In this context, neuromodulatory agents such as gabapentinoids or serotonin–norepinephrine reuptake inhibitors, which reduce neuronal hyperexcitability and enhance descending inhibitory pathways, may represent biologically rational therapeutic options.

Third, careful anatomical evaluation remains critical for identifying patients at risk of structural complications. Classification systems that capture compartmental disease extent, such as the Enzian score, provide valuable information for anticipating surgical complexity and guiding surveillance for complications including ureteral obstruction and infertility.

### Research implications

These findings highlight the need for integrated disease classification frameworks that capture both structural burden and biological activity. Current staging systems quantify anatomical extent but do not measure neuroinflammatory activity or fibrotic remodeling. Conversely, pain scales capture symptom intensity but do not indicate structural risk. Future research should therefore focus on multidimensional phenotyping strategies that combine anatomical staging, symptom profiling, and molecular biomarkers. The transcriptional programs identified in this study also suggest potential therapeutic targets. Modulation of inflammatory signaling, extracellular matrix remodeling, and myofibroblast activation may represent promising approaches for addressing both symptom burden and disease progression.

### Strengths and limitations

This study integrates detailed clinical phenotyping with molecular analysis of lesion tissue, allowing simultaneous evaluation of anatomical disease burden, symptom severity, and transcriptional programs associated with pain. The inclusion of longitudinal reproductive outcomes and surgical variables further strengthens the clinical relevance of our findings. However several limitations should be acknowledged. Transcriptomic analyses were performed in a subset of samples and may not capture the full molecular heterogeneity of endometriosis. Pain assessment relied on patient-reported measures and may be influenced by individual differences in perception and reporting. In addition, the observational design limits causal inference regarding the mechanistic relationships between fibrosis, neuroinflammation, and clinical outcomes.

### Conclusions

Endometriosis severity cannot be adequately conceptualized as a single lesion-based continuum. Instead, disease manifestations arise from the interaction of two partially independent biological programs: a neuroinflammatory axis governing pain and quality-of-life impairment and a fibrotic structural axis determining fertility outcomes, surgical complexity, and risk of organ compromise. Recognizing these distinct yet intersecting dimensions helps explain the longstanding disconnect between lesion burden and symptom severity. Integrating anatomical and biological phenotyping into clinical practice may enable more precise management strategies aimed at alleviating pain while preventing progressive structural complications, including organ-threatening manifestations such as ureteral obstruction and renal failure.

## Data Availability

All data produced in the present study are available upon reasonable request to the authors.

## Figure Legends

**Table 1:**
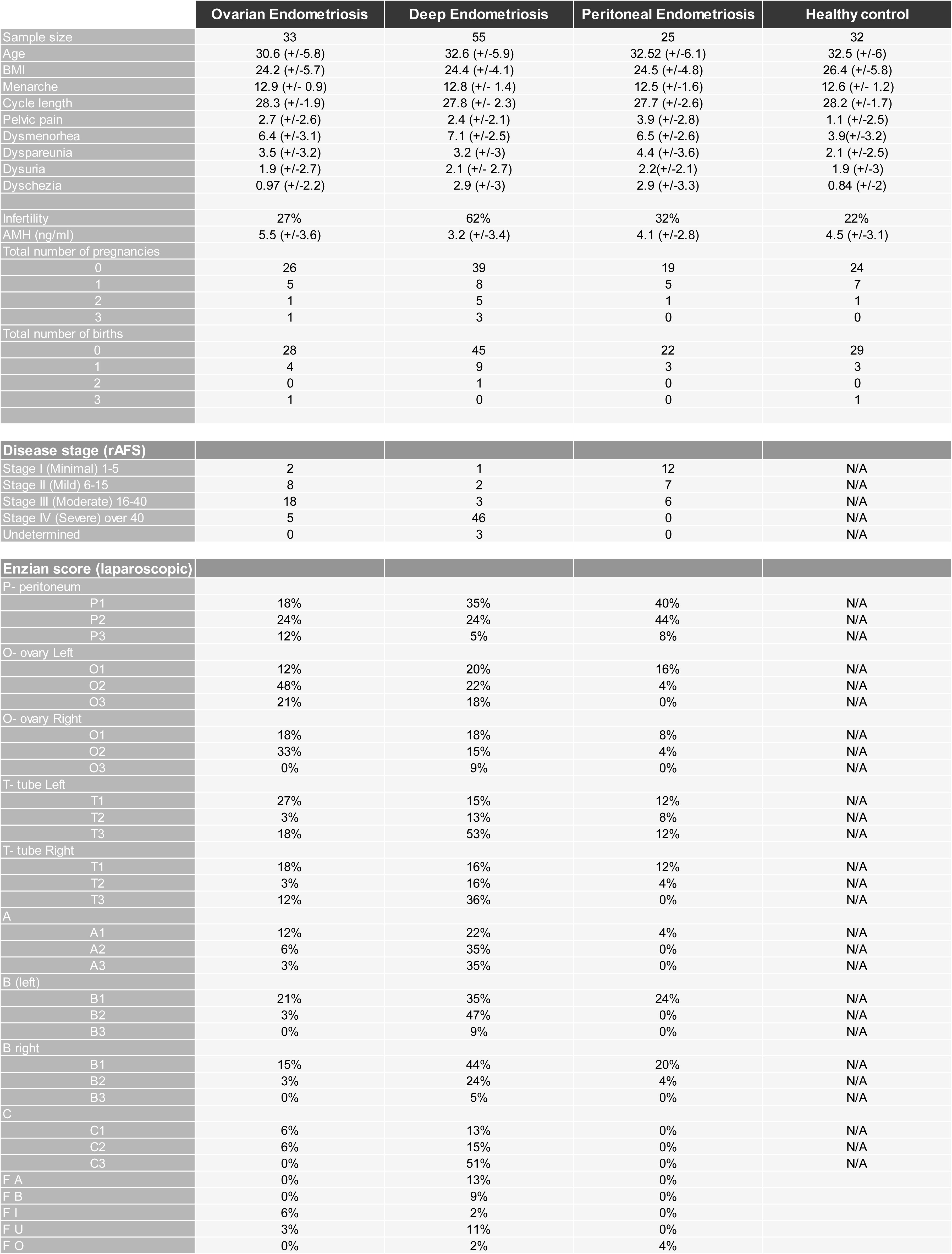
Recruited patients’ characteristics. . Baseline characteristics, symptom severity, reproductive parameters, and anatomical classification across ovarian endometriosis, deep endometriosis, peritoneal endometriosis, and healthy control groups. Continuous variables are presented as mean ± standard deviation. Categorical variables are shown as absolute numbers or percentages, as indicated. Disease stage was classified according to the revised American Society for Reproductive Medicine (rASRM) system. Deep infiltrating lesions were described using the Enzian classification based on laparoscopic findings. AMH, anti-Müllerian hormone; BMI, body mass index; rASRM, revised American Society for Reproductive Medicine.

**Supplementary Table 1.** Predictors of pregnancy at diagnosis and 12-month follow-up. P values, effect differences, and q values for associations between pain domains, structural scores, QoL domains, and pregnancy across OE, DE, and PE. Green highlight indicates significant association between pregnancy and respective characteristics.

|  |  | Ovarian endometriosis |  |  | Deep endometriosis |  |  | Peritoneal endomeriosis |  |  |
| --- | --- | --- | --- | --- | --- | --- | --- | --- | --- | --- |
|  |  | P value | Difference | q value | P value | Difference | q value | P value | Difference | q value |
| Diagnosis | Pelvic pain | 0.017 | -3.267 | 0.431 | 0.845 | 0.175 | 0.945 | 0.939 | 0.167 | 0.980 |
|  | Dysmenorrhea | 0.075 | -2.833 | 0.489 | 0.877 | 0.143 | 0.947 | 0.263 | -2.083 | 0.980 |
|  | Dyspareunia | 0.194 | -2.533 | 0.553 | 0.623 | -0.587 | 0.751 | 0.614 | -1.333 | 0.980 |
|  | Dysuria | 0.701 | -0.667 | 0.985 | 0.278 | -1.349 | 0.580 | 0.776 | -0.500 | 0.980 |
|  | Dyschezia | 0.881 | 0.167 | 0.985 | 0.603 | 0.635 | 0.751 | 0.861 | 0.417 | 0.980 |
|  | Ultrasound Enzian Score | 0.783 | -0.667 | 0.985 | 0.602 | -0.841 | 0.751 | 0.838 | 0.583 | 0.980 |
|  | Stage | 0.297 | -0.400 | 0.596 | 0.137 | -0.413 | 0.389 | 0.432 | 0.500 | 0.980 |
|  | Physical & Functional Impa | 0.913 | -0.433 | 0.985 | 0.594 | -2.333 | 0.751 | 0.706 | 2.750 | 0.980 |
|  | Symptom Burden & Perceiv | 0.665 | -1.600 | 0.985 | 0.734 | -0.857 | 0.851 | 0.881 | 0.917 | 0.980 |
|  | Emotional & Psychological | 0.325 | -2.867 | 0.599 | 0.374 | 2.000 | 0.615 | 0.697 | 1.583 | 0.980 |
|  | Social Identity, Stigma & S | 0.415 | -2.967 | 0.722 | 0.937 | -0.222 | 0.977 | 0.644 | 2.667 | 0.980 |
| 12 Months Follow Up | Pelvic pain | 0.041 | -1.667 | 0.431 | 0.265 | -0.778 | 0.580 | 0.242 | 0.917 | 0.980 |
|  | Dysmenorrhea | 0.078 | -2.000 | 0.489 | 0.064 | -2.556 | 0.224 | 0.088 | -2.833 | 0.980 |
|  | Dyspareunia | 0.867 | -0.200 | 0.985 | 0.151 | -1.333 | 0.393 | 0.657 | -0.833 | 0.980 |
|  | Dysuria | 0.220 | -1.333 | 0.574 | 0.348 | -1.333 | 0.605 | 0.242 | -0.833 | 0.980 |
|  | Dyschezia | 0.855 | 0.133 | 0.985 | 0.597 | -0.556 | 0.751 | 0.467 | 0.833 | 0.980 |
|  | Ultrasound Enzian Score | 0.041 | -3.067 | 0.431 | 0.049 | -2.079 | 0.218 | 0.275 | -0.500 | 0.980 |
|  | Physical & Functional Impa | 0.178 | -1.867 | 0.553 | 0.030 | -7.270 | 0.218 | 0.485 | 3.500 | 0.980 |
|  | Symptom Burden & Perceiv | 0.119 | -1.600 | 0.496 | 0.014 | -5.651 | 0.213 | 0.911 | -0.583 | 0.980 |
|  | Emotional & Psychological | 0.257 | -2.133 | 0.596 | 0.037 | -4.762 | 0.218 | >0.999999 | 0.000 | >0.999999 |
|  | Social Identity, Stigma & S | 0.105 | -2.600 | 0.496 | 0.045 | -5.746 | 0.218 | 0.892 | -0.750 | 0.980 |

**Supplementary Figure 1.**
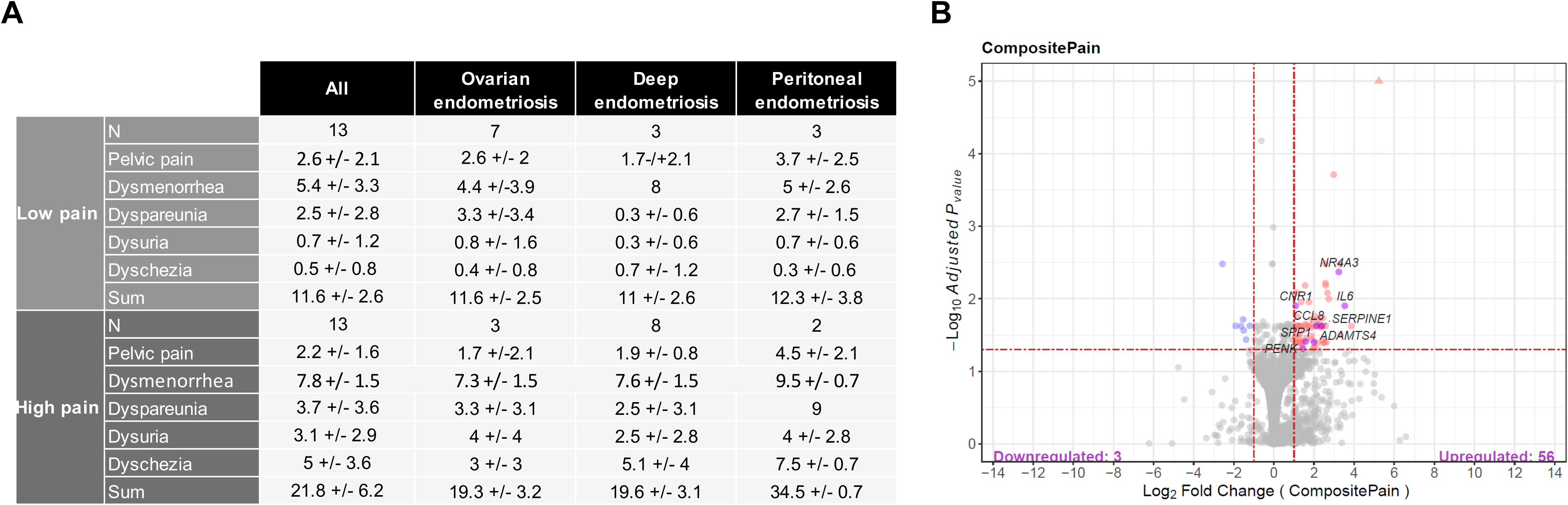
Pain-associated biomarkers. (A) Table showing sample grouping into high and low pain groups and individual pain domain average scores for endometriosis subtypes. (B) Volcano plot depicting the distribution of differentially expressed genes between aggregated endometriosis subtypes in low and high pain groups. The x-axis represents the log_2_ fold change, while the y-axis represents -log_10_(p-value). Each point corresponds to a gene, with significantly upregulated genes shown in red, downregulated genes in blue, and non-significant genes in gray. The dashed horizontal line indicates the significance threshold (p-value < 0.05), and the vertical dashed lines denote the fold-change cut-off (log_2_FC > 1 or < −1). Labeled points indicate key genes of interest.

